# The burden of nosocomial covid-19: results from the Wales multi-centre retrospective observational study of 2518 hospitalised adults

**DOI:** 10.1101/2021.01.18.21249433

**Authors:** Mark J Ponsford, Rhys Jefferies, Chris Davies, Daniel Farewell, Ian R. Humphreys, Stephen Jolles, Sara Fairbairn, Keir Lewis, Daniel Menzies, Amit Benjamin, Favas Thaivalappil, Christopher Williams, Simon Barry

## Abstract

**Objectives:** To define the burden of nosocomial (hospital-acquired) novel pandemic coronavirus (covid-19) infection among adults hospitalised across Wales.

**Design:** Retrospective observational study of adult patients with polymerase chain reaction (PCR)-confirmed SARS-CoV-2 infection between 1^st^ March – 1^st^ July 2020 with a recorded hospital admission within the subsequent 31 days. Outcomes were collected up to 20^th^ November using a standardised online data collection tool.

**Setting:** Service evaluation performed across 18 secondary or tertiary care hospitals.

**Participants:** 4112 admissions with a positive SARS-CoV-2 PCR result between 1^st^ March to 1^st^ July 2020 were screened. Anonymised data from 2518 participants were returned, representing over 60% of adults hospitalised across the nation of Wales.

**Main outcome measures:** The prevalence and outcomes (death, discharge) for nosocomial covid-19, assessed across of a range of possible case definitions.

**Results:** Inpatient mortality rates for nosocomial covid-19 ranged from 38% to 42% and remained consistently higher than participants with community-acquired infection (31% to 35%) across a range of case definitions. Participants with nosocomial-acquired infection were an older, frailer, and multi-morbid population than those with community-acquired infection. Based on the Public Health Wales case definition, 50% of participants had been admitted for 30 days prior to diagnostic testing.

**Conclusions:** This represents the largest assessment of clinical outcomes for patients with nosocomial covid-19 in the UK to date. These findings suggest that inpatient mortality rates from nosocomial-infection are likely higher than previously reported, emphasizing the importance of infection control measures, and supports prioritisation of vaccination for covid-19 negative admissions and trials of post-exposure prophylaxis in inpatient cohorts.

**Trial registration:** This project was approved and sponsored by the Welsh Government, as part of a national audit and quality improvement scheme for patients hospitalised covid-19 across Wales.

**Key Messages:** *What is already known on this topic:* We searched PubMed and ISI Web of Science up until 31-December-2020 for studies reporting on patient outcomes following hospital-acquired infection due to the severe acute respiratory syndrome coronavirus 2 (SARS-CoV-2). We identified a range of case-definitions for hospital-acquired infection, based on timing of diagnostic testing 5 to 15 days following admission. The largest and only multi-centre study concluded individuals with nosocomial infection are at a lower risk of death from SARS-CoV-2 than those infected in the community, however, was performed early in the pandemic and utilised a conservative definition of nosocomial infection.

*What this study adds:* Our multi-centre observational study represents the largest assessment of clinical outcomes for patients with nosocomial covid-19 in the UK to date, and suggests the burden of nosocomial SARS-CoV-2 infection has been underestimated. Nosocomial-infection occurred in older, frailer, and multi-morbid individuals, and was consistently associated with greater inpatient mortality than amongst those who were infected in the community across a spectrum of case-definitions. Our findings support implementation of enhanced infection control measures to reduce this burden during future waves, especially given the recent emergence of novel viral variants with enhanced transmissibility. Furthermore, roughly half of the patients meeting the Public Health Wales definition of definite nosocomial SARS-CoV-2 infection had been admitted for 30 days prior to diagnosis, highlighting a potential window of opportunity for inpatient pre-exposure and/or post-exposure prophylaxis.

## Introduction

Health-care-associated infections represent an enduring and serious threat to patient safety in hospital and healthcare facilities ^1,2^ and have been estimated to cost the National Health Service £1 billion every year.^3^ The transmission of respiratory viruses such as influenza in the healthcare environment are a well recognised cause significant morbidity and mortality at the individual patient level,^4^ however little is known regarding the significance of in-hospital (nosocomial) transmission of the novel pandemic coronavirus, SARS-CoV-2. Such information is essential to assess the effectiveness of recent infection control measure guidance.^5^ A national report published by the Healthcare Safety Investigation Branch on 29th October 2020 stated: “*At the time of writing, there was no publicly available data to provide evidence of the scale of nosocomial of covid-19 in the NHS*”.^6^ In addition, acknowledgement of risk is timely to inform prioritisation of public health measures such as vaccination of healthcare workers and vulnerable populations, and decisions regarding continuation of routine NHS service provision.

We identified limited studies reporting on the prevalence and outcomes of medical patients’ hospital-acquired covid-19 within the UK,^7-14^ employing a range of case definitions and study design – restricting generalisability. The largest and only multi-centre cohort study was performed the by COPE (COVID-19 in Older PEople study) investigators, describing outcomes in 1564 patients admitted with polymerase chain reaction (PCR)-confirmed SARS-CoV-2 infection across 11 hospitals (including 4 in Wales) between 27^th^ February until 28^th^ April 2020.^8^ Applying a probabilistic case definition of nosocomial infection (admitted for at least 15 days before a SARS-CoV-2 diagnosis), Carter et al. estimated 12.5% contracted covid-19 in hospital. This increased to 23.0% if diagnoses made between 5 and 14 days of admission were included. Mortality in the nosocomial patient group appeared comparable to those with likely community-acquired infection (27.0% and 27.2%, respectively), despite the older age and higher frailty of these patients.^8^ Applying a similar case definition, Taylor et al found probable nosocomial cases represented 11.3% of confirmed cases within a major London trust admitted between 6^th^ March to 12 April 2020; mortality was unreported in this study.^7^ These reports preceded recognition of high-profile nosocomial outbreaks across several UK hospitals.^15,16^ Thus, reliable estimates of the true impact of hospital-acquired covid-19 infection remain hampered by a paucity of publicly-available data at national and regional levels.^17^

Here, we update assessment of the relative burden of community-relative to nosocomial-acquired SARS-CoV-2 infection, utilising anonymised patient- and hospital-level data collected via the *National Pathway for Managing Covid-19 Infections in Secondary Care in Wales* initiative. Our primary aim was to describe inpatient case load and mortality across the nation of Wales in the context of probable infection source. Our secondary aim was to examine the significance of commonly used differing case definitions and statistical methodologies when interpreting these findings. Together these findings suggest that mortality from nosocomial infection may be higher than previously reported, emphasizing the importance of infection control measures to protect vulnerable inpatient cohorts, and directing prioritisation of pre- and post-exposure prophylactic measures.

## Methods

### Study design

Data were obtained as part of the National Pathway for Managing Covid-19 Infections in Secondary Care in Wales initiative (www.covid-19hospitalguideline.wales.nhs.uk), which collects patient- and site-level outcomes through the use of an online data collection tool, supporting guideline development and subsequent audit cycles.

### Setting

In each hospital in Wales a study lead was identified, who coordinated data collection. All 18 major hospitals in Wales that admitted patients through emergency departments or medical assessment units were included (Supplementary Table 1).

### Participants

Positive SARS-CoV-2 polymerase chain reaction (PCR) results recorded between 1^st^ March 2020 and 1st July 2020 with a record of hospital admission were obtained from Public Health Wales and used to identify patients for retrospective notes review information by local clinical teams.

### Data collection

Demographic variables with prognostic significance were collected as mandatory data fields for the index admission using an online tool including: date of positive PCR swab collection, dates of admission and discharge (when complete), patient age, sex, comorbidity count, and outcome (death or discharge). Supplementary fields were also collected, as described in the text.

### Outcomes

The primary outcome for analysis was descriptive in nature, with comparison of all-cause mortality rate and cohort demographics based upon probable origin of SARS-CoV-2 infection. Secondary analyses based on time to discharge or death were also investigated.

### Missing data

Missing data were included in tables and descriptive analyses, allowing denominators to remain consistent in calculations. Imputation of missing data was not performed.

### Bias

Notes were selected randomly by site audit/record departments, to minimise potential for systematic bias prior to data entry.

### Quantitative variables

Frailty was assessed using the pre-admission Clinical Frailty Scale (CFS), as recommended by the National Institute for Clinical Excellence.^18^ For the purposes of the analyses, CFS scores were grouped into clinically meaningful groups: 1-4, 5–6, and 7–9. Welsh Index of Multiple of Deprivation (WIMD) was derived from the patients’ post-code and evaluated by quartiles.

### Statistical methods

Baseline demographic and clinical characteristics assessed at admission were grouped by outcome and probable infection origin, based on a) clinician-recorded case definition, b) standardised case definitions (Supplementary Table 2). Continuous variables are presented as median [inter-quartile range, IQR] and categorical variables as n (%), unless otherwise stated. Time-to-event data were analysed using cumulative incidence curves, acknowledging the competing risk of death or discharge. We used time in hospital following a covid-19 diagnosis as a standard measure to avoid introducing survivorship-bias, defining day 0 of the at-risk period as the most recent of either admission date or SARS-CoV-2 diagnostic testing.

Secondary statistical significance testing was performed according to the data encountered: For univariate analysis of categorical data, such as sex, Fisher’s exact or chi-square testing was performed, comparing observed numbers for NC cases with numbers predicted by the ratio within the CAC sub-group. For continuous data, Welch’s t-tests were used if the assumptions of normality were met; otherwise non-parametric Mann-Whitney U tests were employed. Multi-level logistic regression was used to calculate odds ratios (ORs) and 95% Confidence Intervals (CI), including factors that occurred before the outcome of interest to preserve causal order. Two-sided statistical significance was set at p <0.05. Analysis were performed using R version 4.0.2 in RStudio (Version 1.3.959, R Foundation, Vienna, Austria) with packages including *tidyverse*, and *survival* and GraphPad Prism (version 6.07).

Sensitivity analyses were performed to assess the effect of varying the clinical case definition for nosocomial-acquired infection, and to minimise potential survivorship bias associated with previously-used case definitions.

This project constitutes the evaluation/audit arm of the National Pathway for Managing covid-19 Infections in Secondary Care in Wales and was approved by Welsh Government. Information governance policies on data protection were followed to record data securely at each site. Anonymised data handling and storage was supported by The Institute of Clinical Science and Technology, Cardiff UK.

Patient and public involvement: These data were generated as part of a rapid appraisal process and as such patients were not involved in the setting of the research question or interpretation of the study. Findings will be disseminated via the www.covid-19hospitalguideline.wales.nhs.uktool.

## Results

### Participants

We identified 6005 SARS-CoV-2 positive results with a location in hospital, taken between 1/3/2020 and 1/7/2020 inclusive, of which 4112 were individual cases. These were screened from which clinical case information on 2594 case records were inputted (Supplementary Figure 1: Study flowchart). Admission dates ranged from 25^th^ March 2019, to 3^rd^ July 2020, with the last recorded discharge on 26^th^ October 2020. A total of 76 individuals were excluded from the study after data screening due to missing primary data fields. Reasons for exclusion were: age <18 years or not stated (n=27), admission date missing or incorrectly entered (n=15), date of PCR diagnosis missing or outside of study period (n=16), and discharge date incorrectly entered (n=3). In addition, 15 cases where the first PCR result exceeded 31 days of admission or discharge were excluded. This left 2518 patients for analysis with complete case records for primary fields, representing approximately 63% of the total adult population hospitalised with covid-19 within the nation of Wales. For 10 of the 18 participating sites ≥ 80% coverage was achieved. Completeness of supplementary data fields was more varied, with WIMD score available in 95.1% of cases, admission source in 2372 (93.8%), inpatient ceiling of care decisions in 1976 (78.2%), and CFS in 1316 cases (52.1%).

### Descriptive data

Admission features are summarised by outcome in Table 1. The cohort comprised a relatively elderly population with a median age of 74 years [IQR 62.5-85.5; range 19-99]. Overall, more men (n= 1369, 54.4%) than women (n=1149, 45.6%) were admitted to hospital with a diagnosis of covid-19, although this trend was reversed within the under 45 and over 85-year age groups (Table 1; Supplementary Table 3). A deprivation gradient was also apparent, with individuals from the most-deprived (lowest) WIMD quartile over-represented within admissions (31.1%) relative to those in the least deprived quartile (18.9%, Chi-squared test: p <0.0001).

**Table 1:**
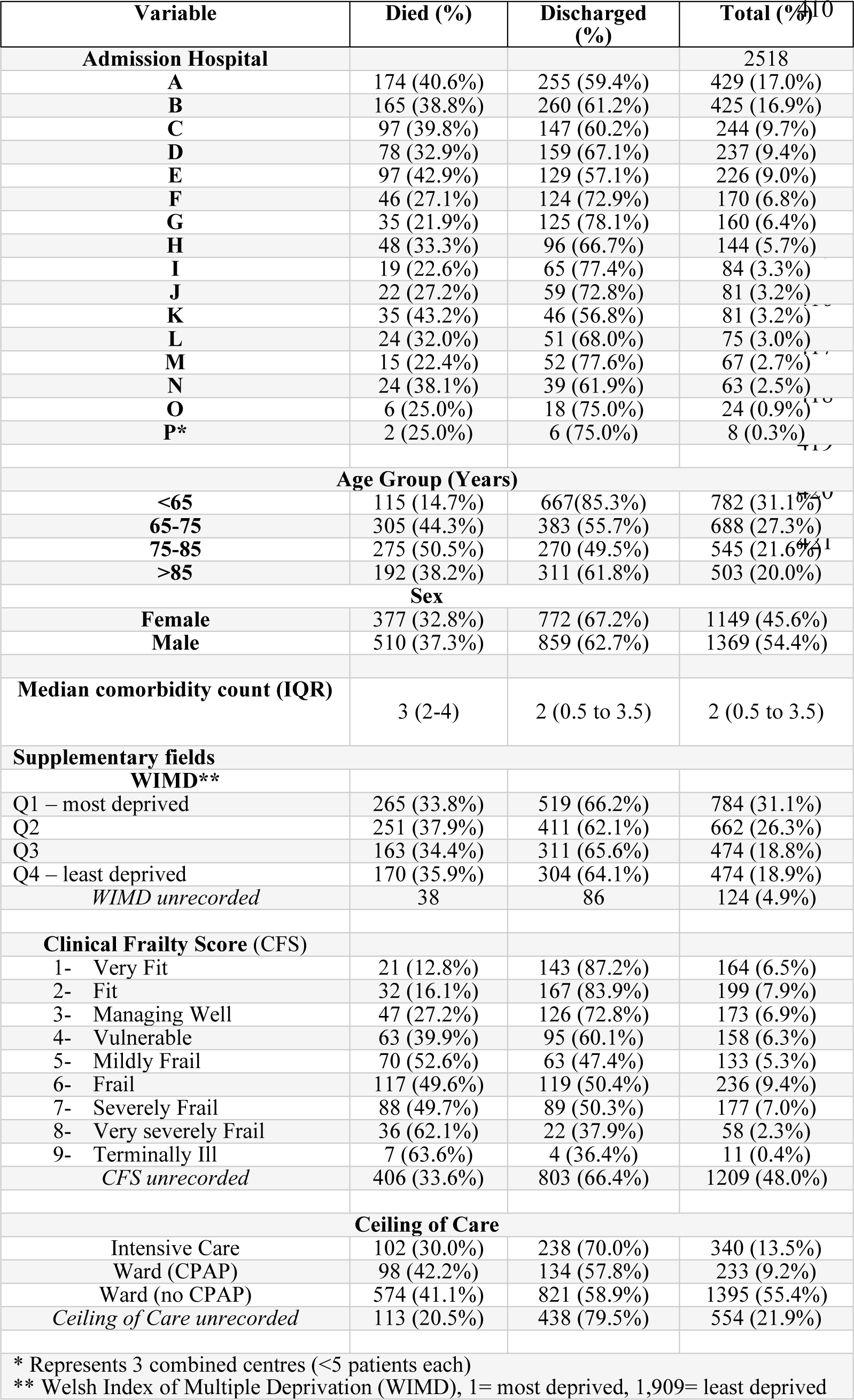
Demographics and clinical features at presentation

### Prevalence and mortality rates by clinician recorded admission source

To investigate the impact of covid-19 infection origin on patient outcomes, we first examined mortality by clinician-defined admission source, available in 2372 cases (93.8%); shown Table 2. Hospital-acquired covid-19 was documented in 437 cases (17.3% of cohort, 37.8% mortality), comparable to mortality following ambulance presentation (40.5%). Walk-in and GP referrals together accounted for 20.4% of the cohort and had the lowest estimated inpatient mortality rate (17.1-23.2%), comparable to patients without a recorded admission source (26.3%). The small number of patients admitted from care or nursing homes (n=52) showed an inpatient mortality rate of 46.2%.

**Table 2:** Patient outcomes by clinician-defined admission source

| <u>Admission Source</u> | <u>Died (%)</u> | <u>Discharged (%)</u> | <u>Total cases (%)</u> |
| --- | --- | --- | --- |
| **Ambulance** | 554 (40.6%) | 810 (59.4%) | 1364 (54.2%) |
| **GP Practice** | 61 (23.4%) | 200 (76.6%) | 261 (10.4%) |
| **Walk In** | 43 (17.1%) | 209 (82.9%) | 252 (10.0%) |
| **Acquired in Hospital** | 164 (37.7%) | 271 (62.3%) | 435 (17.3%) |
| **Other/not defined** | 41 (26.6%) | 113 (73.4%) | 154 (6.1%) |
| **Care Home** | 24 (46.2%) | 28 (53.8%) | 52 (2.1%) |

### Nosocomial covid-19 is associated with greater mortality than community-acquired cases across a range of standard case definitions

We next applied a definition of nosocomial covid-19 reported by the COPE-study investigators and used by Public Health Wales,^8^ based upon an interval exceeding 14 days between admission and SARS-CoV-2 diagnostic testing (Supplementary Table 1). We identified 413 nosocomial cases, representing 16.4% of the cohort. Most patients (n=1610, 63.9 %) were diagnosed by PCR sampling conducted prior to or within the first five days of admission date and were defined as community-acquired; 456 patients (18.1% of total cases) had a positive PCR taken between 5 and 14 days after admission. By end of the study period, 39.2% of patients with nosocomial-infection had died, compared to a case fatality rate of 31.6% in community-acquired covid-19 cases. This observation of greater mortality amongst NC patients proved generalisable across the majority of admission sites (Figure 1). The median time from SARS-CoV-2 diagnosis to discharge in community-acquired covid-19 cases was 7 days (IQR: 3.0-15.0), compared to 17 days (IQR: 7.0-38.0) in patients with nosocomial-infection. Using this definition, nosocomial cases had been admitted for a median of 30 days prior to diagnosis (IQR: 15.0-62.5), with 48 patients admitted for over 100 days. The median overall length of stay for patients with nosocomial infection was 60 days (IQR: 34.0-98.5).

**Figure 1:**
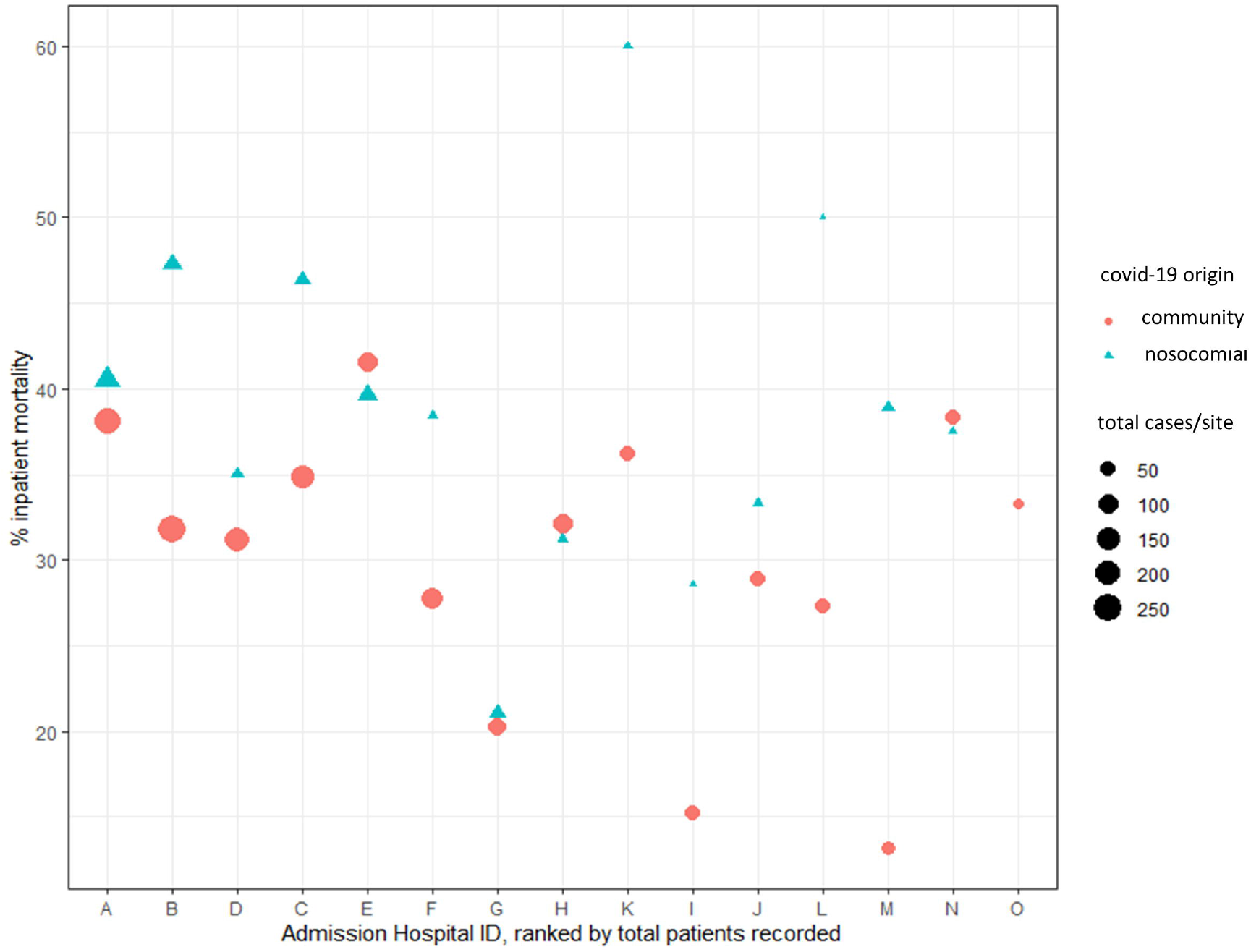
Inpatient mortality rates by admission hospital sites Scatter plot showing inpatient mortality rates for patients with community-acquired covid-19 (circles); and nosocomial covid-19 (triangles) by individual sites, with hospitals arranged by decreasing overall case load are plotted from the left. For 11/15 sites, inpatient mortality rates for nosocomial cases exceeds that of community acquired cases. Individual sites with fewer than 5 cases were excluded from analysis.

As 95% of patients will display symptoms between 2 to 11.5 days of exposure,^19^ the COPE-study definition represents a conservative estimate of the burden of nosocomial-cases infection. We therefore considered the effect on mortality and case numbers by varying the case definition across the incubation period, taking admission as the earliest potential nosocomial exposure (Supplementary Table 4). As shown in Figure 2A, inpatient mortality rates for nosocomial covid-19 ranged from 38% to 42% and remained consistently greater than in patients with community-acquired infection (31% to 34%). By contrast, varying the case definition resulted in significant changes in case numbers and deaths (Figure 2B/C). Applying the Public Health England case definition (diagnosis of SARS-CoV-2 made >7 days following admission) identified 727 admissions and 301 deaths. This rose to 830 cases and 342 deaths when patients with a date of PCR testing >5 days following admission was used to diagnose nosocomial-covid-19.^20^ Together, these findings indicate that whilst changing the case definition for nosocomial-covid-19 has limited effect on case fatality rates, it significantly alters the number of reported cases of nosocomial infection and total mortality.

**Figure 2:**
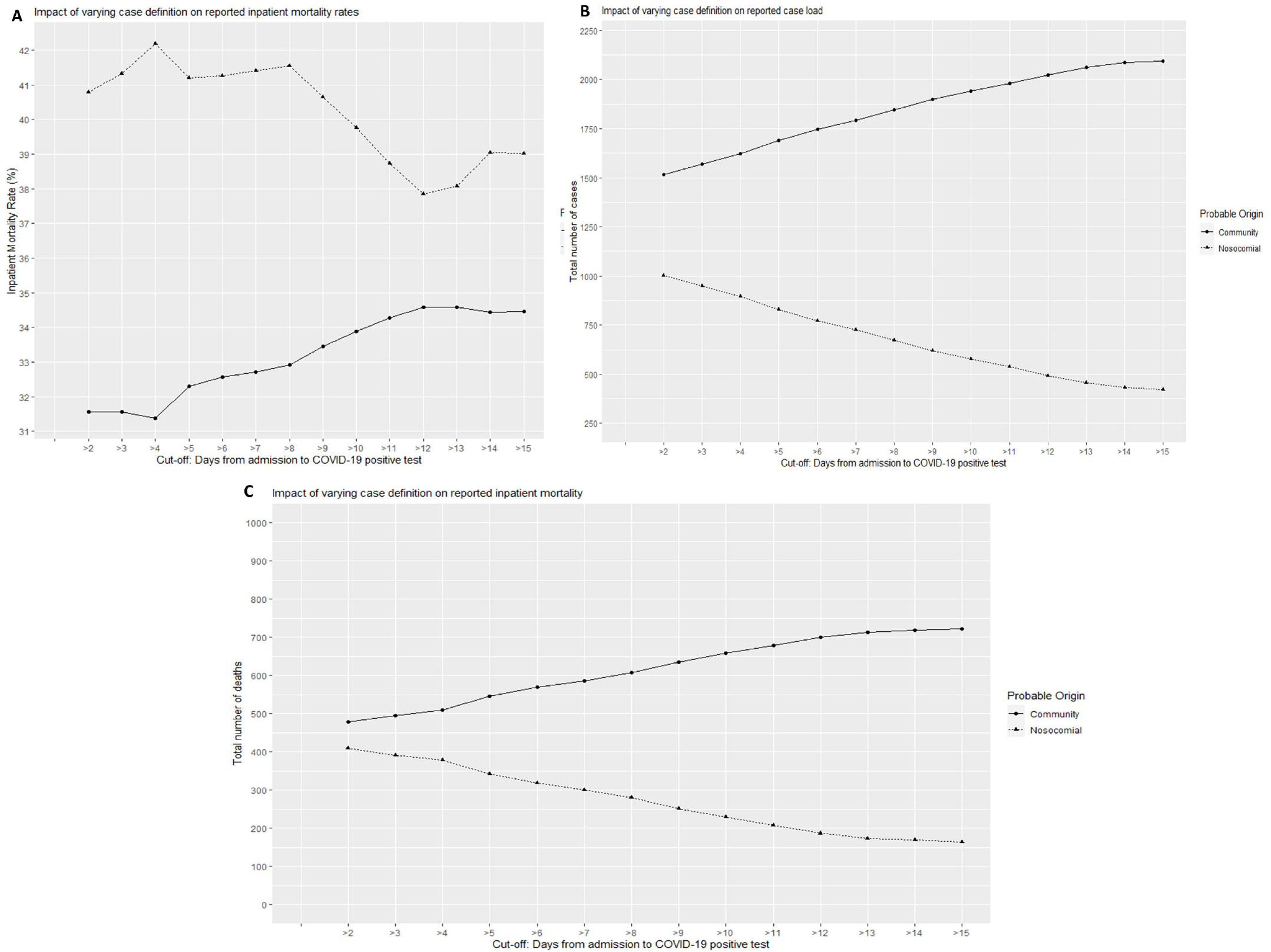
Effect of varying case definitions on prevalence and crude mortality rate for community- and nosocomial-SARS-CoV-2 infection. Sensitivity analyses considering the effect on inpatient mortality rate (A), total case numbers (B), and overall mortality burden (C) by varying the case definition across the incubation period, taking admission as the earliest potential nosocomial exposure.

### Competing Risk analysis provides differing conclusions to Kaplan Meier

Kaplan Meier survival analysis has been utilised by several recent groups to compare outcomes for hospital and community-acquired covid-19 in UK hospitals to date.^8,9^ However, the differing discharge kinetics observed in patients with nosocomial- and community-acquired covid-19 led us to question the suitability of this statistical methodology without consideration of the competing risks of discharge to death. As shown in Figure 3, nosocomial-infection is associated with a greater cumulative incidence of inpatient mortality than community-acquired, in line with the crude case inpatient mortality rates described. This contrast with Kaplan Meier analysis (Supplementary Figure 2), confirming the potential for conflicting interpretations of the same data set when employing different statistical methodologies.

**Figure 3:**
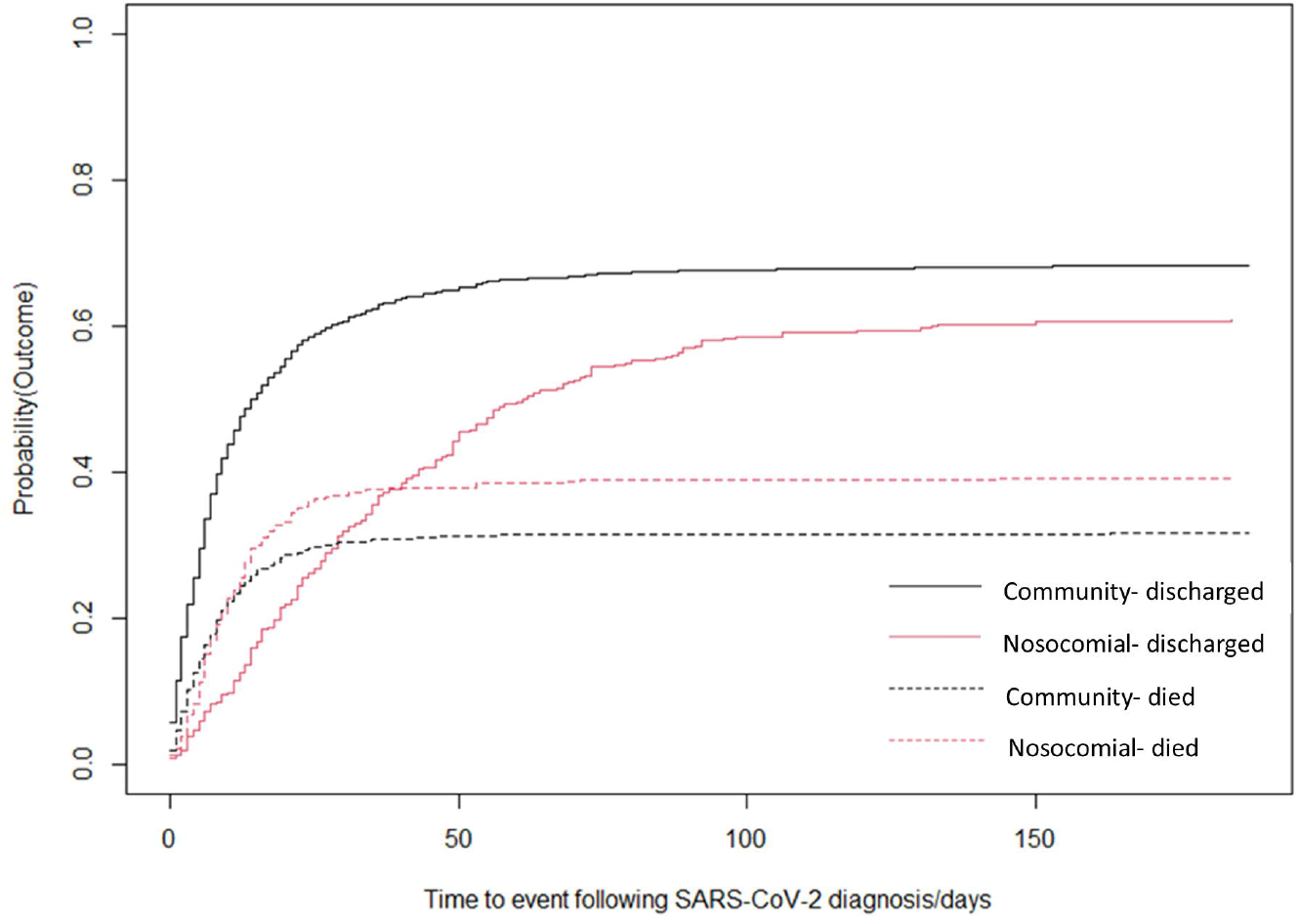
Competing risk analysis plot of nosocomial and community infection outcomes of covid-19 patients. Time to event analysis cumulative incidence analysis for the competing risks of discharge and diagnosis, using the time from SARS-CoV-2 diagnosis. covid-19 origin is assigned by the commonly used case definition, as outlined by Carter et al ^8^, nosocomial shown in red, community-acquired covid-19-shown in black. Dotted lines: cumulative incidence of death, continuous lines: cumulative incidence of discharge on probability scale. To deal with potential survivorship bias introduced by including CAC patients tested prior to admission (who cannot reach discharge or death until admission), Day 0 is defined as the more recent of day of admission or date of diagnostic SARS-CoV-2 testing.

### Patients with probable nosocomial COVID-19 infection are a more vulnerable population than community acquired cases

We hypothesized that patients with nosocomial-covid-19 might represent a more vulnerable population, a finding suggested by previous groups, but potentially confounded by introduction of selection bias due to the requirement for a prolonged inpatient admission period within the case definition of nosocomial-SARS-CoV-2 infection. To address this, we explored demographic and clinical characteristics in patients based on a diagnostic cut-off of 2 days post-admission, a definition commonly used for hospital-acquired bacterial pneumonia. Individuals with a positive SARS-CoV-2 test taken >2 days following admission were typically older and frailer, with a greater median CFS of 6 (IQR 4.5-7.5), compared to 3 (IQR 1-5) in those with a positive SARS-CoV-2 test taken up to 2 days following admission. Marked differences in multi-morbidity were evident, with 35.4% of “nosocomial” patients having at least 4 comorbidities compared to 26.6% of “community-acquired” patients (Supplementary Table 5). Together, this supports the hypothesis that age, frailty, and multi-morbidity are factors associated with hospital-acquired covid-19, and not simply a reflection of the diagnostic definition.

Given the apparent vulnerability of the patient cohort developing nosocomial infection, we questioned if ceiling of care decisions might impact upon patient outcomes. Returning to the COPE-study definition, ceiling of care decisions were recorded in 1249 (77.6%) of community-acquired and 322 (78.0 %) of nosocomial-acquired covid-19. A ward-based ceiling of care (including consideration of CPAP or NIV) was documented in 75.3% of nosocomial cases, compared to 58.8% of community-acquired cases (Fisher’s Exact Test p<0.0001, OR: 2.13, 95% CI: 1.58 to 2.87). Put another way, the odds of a ward level ceiling of care being documented was twice as great in patients with nosocomial-acquired compared to community-acquired covid-19. To determine the relative importance of this as a predictor of adverse outcome, we performed logistic regression analysis with the binomial outcome of death or discharge in nosocomial-acquired cases. In univariate analysis, older age and male sex were both associated with increased odds of death; OR 1.02 (95% CI: 1.03 to 1.05) and OR 1.56 (95% CI: 1.05 to 2.33), respectively. Recorded ceiling of care at ward level was significantly associated with an increased risk of mortality: OR 3.20 (95% CI: 1.88 to 5.43) when considered alone, but not following adjustment for age, sex, index of multiple deprivation and comorbidities OR 1.39 (95% CI: 0.572 to 3.38). Taken together, these observations confirmed our expectation that patient with nosocomial-acquired SARS-CoV-2 represented a more vulnerable cohort, and as such were less likely to be managed using more invasive care such as intubation or CPAP. However, we found no evidence that a ward-level ceiling of care was an independent predictor of mortality.

### Monthly prevalence of nosocomial covid-19

We determined monthly trends for the frequency of total and new nosocomial diagnoses (Supplementary Table 6). Between March and April 2020, the number of cases meeting the COPE definition for nosocomial-infection across Wales increased from 97 to 216, paralleling the rise in total covid-19 admissions. Nosocomial-infection represented 21.0% of new covid-19 diagnoses during May (73/347 total cases), falling slightly to 18.5% in June. Similar nosocomial infection rates were observed based on clinician-recorded nosocomial diagnoses, with a monthly prevalence between March to June of 17.8% and 19.3%.

## Discussion

Understanding the risk and burden of covid-19 infection within healthcare environments is essential to direct risk mitigation strategies for the immediate future. In this work, we analysed a total of 2,518 patients with PCR-proven covid-19 infection admitted to hospitals across Wales. In contrast to previous studies,^8^ reported earlier in the pandemic, inpatient mortality rates were consistently greater in those who likely acquired the virus in the context of hospital admission (37.9-42.2%), compared to community-acquired infection (31.4-34.6%) across a range of case definitions. Such heightened mortality rate estimates are in line with single centre reports from a major London teaching hospital where definitely or probably hospital-acquired cases had a fatality rate of 36%.^13^ Similarly, the monthly prevalence estimates found in our study are also in line with the 20% modelled for a typical UK hospital.^21^

Our study is also the first to investigate the impact of applying a range of case definitions when quantifying the burden of hospital-acquired covid-19. By adopting a clinical case definition based on the median incubation period for SARS-CoV-2 of 5 days,^20^ we identified 14.2% additional cases, and 13.6% more deaths attributable to nosocomial covid-19 than with the >7-day threshold currently used by Public Health England. This increased burden is likely an overestimate but may be relevant given that even low-risk clinical assessment areas may contain patients with SARS-CoV-2.^22^ Thus, in the absence of sufficient isolation cubicles for patients undergoing assessment and adequately sensitive rapid turn-around diagnostic testing, the risk of patient-to-patient transmission is likely to persist despite infection control practices of cohorting and universal screening. We also confirm that patients with hospital-acquired covid-19 appear a disproportionately vulnerable group, seemingly independent of diagnostic delay from admission. Advanced age, frailty, and multi-morbidity are well-recognised risk factors for adverse outcome in community-acquired covid-19.^23,24^ We suggest increased frailty is also likely to be causally associated with an increased personal care requirement and thus predispose to nosocomial exposure of SARS-CoV-2 from healthcare workers, a conclusion supported by recent whole-genome sequencing of nosocomial outbreaks.^14^ As highlighted in a recent evaluation of serial PCR screening in healthcare workers, 3% of those attending work tested positive for SARS-CoV-2.^25^ Serological studies suggest the rate of staff infection is far higher on wards with known nosocomial outbreaks, in studies conducted in hospitals not undertaking staff screening.^26^ Finally, we also considered the impact of ceiling of care on outcome, as few other UK multi-centre studies have explored this. Importantly, we found no evidence that ceiling of care determined outcome once clinical risk factors were accounted for, a finding that may be reassuring for patients and families when discussing this issue.

This study is primarily limited by its retrospective nature, which might predispose to ascertainment bias (for instance, notes being more readily available for patients who have died than awaiting discharge). This risk is common to other multi-centre studies,^8,23,24^ and sites adopted random notes retrieval to minimise this. We further sought to mitigate this by extending the data collection period well beyond the end of the first wave, enabling capture of late discharges and including nosocomial outbreaks. Record entry by a body of motivated clinicians followed a standardised online tool with a limited core requirement, resulting in minimal missing core data. As a result, coverage exceeded 60% of hospital admissions with covid-19 across the nation of Wales (and ≥80% in 10 of 18 participating sites). This compares favourably to recent descriptive studies.^8,23,24^ We recognise that our findings represent crude inpatient mortality rate estimates, based on all-cause mortality. As the total number of patients at risk of infection was unknown, they should not be used to infer the risk of acquiring SARS-CoV-2 within hospital. Future studies making use of linked datasets are suggested to estimate excess mortality associated with nosocomial infection and identify routes of transmission are required. However, with the NHS facing winter-pressures and further surges in SARS-CoV-2 community circulation rates, our findings serve as a timely reminder as to the dangers faced by patients within hospital as well as those within care homes or shielding within the community. This concern is heightened by the spread of a novel antigenic variant with evidence for enhanced transmissibility ^27^

Together, our results highlight the burden associated with extremely infectious agents within the healthcare environment. Although the incidence of nosocomial covid-19 cases decreased significantly following the peak of community-acquired cases, the importance of enhanced infection control measures cannot be overstated.^28^ Indeed, current Public Health Wales data indicates an active rise in probable hospital-acquired case numbers, mirroring the first wave.^29^ We highlight the opportunity for pre-exposure and post-exposure prophylactic measures, including vaccination and/or enrolment in clinical trials employing pharmaceutical and biological antiviral agents such as antibody therapy.^30, 31^ This follows the observation that many of those dying with hospital-acquired covid-19 had typically been in hospital for over a month prior to diagnosis of infection. Inpatient vaccination has long been considered a missed opportunity for improving influenza vaccine uptake.^32^ Finally, we caution that research is urgently required to assess the efficacy of vaccination within vulnerable groups of individuals within hospital and care homes, who are likely to differ from those enrolled in initial trials in age, frailty, and immunocompetence.^33^

## Data Availability

Data sharing will be considered by the management group upon written request to the corresponding author. De-identified participant data or other prespecified data will be subject to a written proposal and signed data sharing agreement.

## Author contributions

SMB conceived the study and worked with RJ and CD to create the covid-19 guideline and the database. MJP led data analysis supported by DF and wrote the first draft supervised by SB1, DF, SJ, and IH. CD leads the Institute for Clinical Science and technology which created the digital interventions. MJP, RJ, CD, and SMB have verified the underlying data and act as guarantors. All authors contributed scientific and clinical guidance regarding study design, data collection, and have reviewed the final draft. The corresponding author attests that all listed authors meet authorship criteria and that no others meeting the criteria have been omitted.

## Acknowledgements

This work uses data collected by the NHS and Public Health Wales as part of the routine care and support for patients admitted with covid-19. We are extremely grateful to the frontline NHS junior doctors for their commitment to inputting data from across Wales. We further acknowledge Gareth Davies and Bea Addison from ICST for (Institute of Clinical Science and Technology) for database management and coordinating the data collection.

