## Supplementary for "The burden of nosocomial covid-19: results from the Wales multi-centre retrospective observational study of 2518 hospitalised adults"

Supplementary Table 1: List of participating centres

| | |
|---|---|
| Aneurin Bevan Health Board | Nevill Hall Hospital, Royal Gwent Hospital and Ysbyty Ystrad Fawr. |
| Betsi Cadwalladr Health Board | Glan Clwyd Hospital, Wrexham Maelor Hospital and Ysbyty Gwynedd (Bangor). |
| Cardiff and Vale Health Board | University Hospital Llandough and University Hospital of Wales. |
| Cwm Taf Health Board | Prince Charles Hospital, Princess of Wales Hospital and Royal Glamorgan Hospital. |
| Hywel Dda Health Board | Bronglais Hospital, Glangwili General Hospital, Prince Phillip Hospital and Withybush General Hospital. |
| Swansea Bay Health Board | Morriston Hospital, Singleton Hospital and Neath Port Talbot Hospital. |

All hospitals delivered urgent and emergency care to patients diagnosed with covid-19.

Supplementary Table 2: Commonly used case definitions for probable covid-19 origin

| | **Case definition** | **Probable covid-19 origin** |
|---|---|---|
| COPE study investigators [1] | Positive SARS-CoV-2 test taken prior to or within first 5 days of admission | Community-acquired covid-19 (“CAC”) |
| | Positive SARS-CoV-2 test taken between 5-14 days after admission | Probable Community-acquired |
| | Positive SARS-CoV-2 test taken more than 14 days after hospital admission (*patient required to remain an inpatient on date of swab sampling*) | Hospital-acquired (Nosocomial- “NC”) |
| Public Health England | Positive SARS-CoV-2 test taken after 7 days of hospital admission | Probable Nosocomial covid-19 |
| Public Health Wales | Positive SARS-CoV-2 test taken an no hospital admission within 28 days or within 2 days of hospital admission | Community onset |
| | Positive SARS-CoV-2 test taken more than 2 days and less than 8 days from hospital admission | Indeterminate hospital onset |
| | Positive SARS-CoV-2 test taken more than 7 days and less than 15 days from hospital admission | Probable hospital onset |
| | Positive SARS-CoV-2 test taken more 14 days from hospital admission | Definite hospital onset |

[1] Carter B, Collins JT, Barlow-Pay F, et al. Nosocomial COVID-19 infection: examining the risk of mortality. The COPE-Nosocomial Study (COVID in Older PEople). *Journal of Hospital Infection* 2020; **106**(2): 376-84.

Supplementary Figure 1: Study flowchart

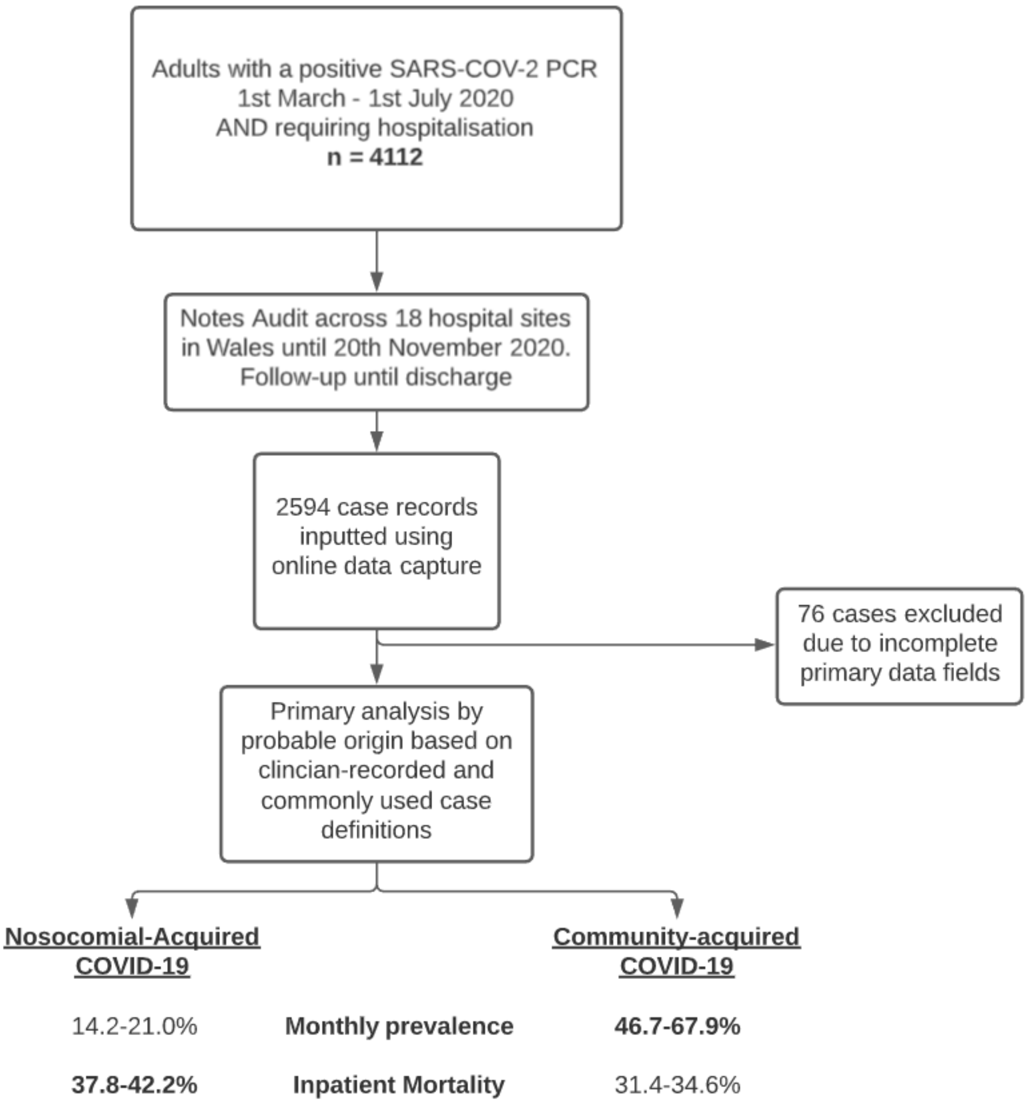

Supplementary Table 3: Admission characteristics stratified by age and sex

| Age Group (years) | Female | % Female | Male | % Male | Total cases | % Total Cases |
|---|---|---|---|---|---|---|
| **18-44** | 98 | 54.1 | 83 | 45.9 | 181 | 7.2% |
| **45-54** | 106 | 46.1 | 124 | 53.9 | 230 | 9.1% |
| **55-64** | 144 | 38.8 | 227 | 61.2 | 371 | 14.7% |
| **65-74** | 196 | 39.0 | 307 | 61.0 | 503 | 20.0% |
| **75-84** | 291 | 42.3 | 397 | 57.7 | 688 | 27.3% |
| **≥85** | 314 | 57.6 | 231 | 42.4 | 545 | 21.6% |

### Supplementary Table 4: Effect of varying diagnostic cut-off on burden of nosocomial infection

Sensitivity analysis to explore the effect of theoretical and current case definitions across the reported incubation period of SARS-CoV-2 virus. Diagnostic cut-off refers to the number of days elapsed between admission and a positive SARS-CoV-2 test being taken. An interval greater than this threshold is used to define nosocomial covid-19, and an interval less than the threshold

| DIAGNOSTIC CUT-OFF | PROBABLE ORIGIN | DIED | DISCHARGED | TOTAL | MORTALITY (%) |
|---|---|---|---|---|---|
| **>2** | Nosocomial | 409 | 594 | 1003 | 40.78 |
| **≤2** | Community | 478 | 1037 | 1515 | 31.55 |
| **>3** | Nosocomial | 392 | 557 | 949 | 41.31 |
| **≤3** | Community | 495 | 1074 | 1569 | 31.55 |
| **>4** | Nosocomial | 378 | 518 | 896 | 42.19 |
| **≤4** | Community | 509 | 1113 | 1622 | 31.38 |
| **>5** | Nosocomial | 342 | 488 | 830 | 41.20 |
| **≤5** | Community | 545 | 1143 | 1688 | 32.29 |
| **>6** | Nosocomial | 318 | 453 | 771 | 41.25 |
| **≤6** | Community | 569 | 1178 | 1747 | 32.57 |
| **>7** | Nosocomial | 301 | 426 | 727 | 41.40 |
| **≤7** | Community | 586 | 1205 | 1791 | 32.72 |
| **>8** | Nosocomial | 280 | 394 | 674 | 41.54 |
| **≤8** | Community | 607 | 1237 | 1844 | 32.92 |
| **>9** | Nosocomial | 252 | 368 | 620 | 40.65 |
| **≤9** | Community | 635 | 1263 | 1898 | 33.46 |
| **>10** | Nosocomial | 229 | 347 | 576 | 39.76 |
| **≤10** | Community | 658 | 1284 | 1942 | 33.88 |
| **>11** | Nosocomial | 208 | 329 | 537 | 38.73 |
| **≤11** | Community | 679 | 1302 | 1981 | 34.28 |
| **>12** | Nosocomial | 187 | 307 | 494 | 37.85 |
| **≤12** | Community | 700 | 1324 | 2024 | 34.58 |
| **>13** | Nosocomial | 174 | 283 | 457 | 38.07 |
| **≤13** | Community | 713 | 1348 | 2061 | 34.59 |
| **>14** | Nosocomial | 169 | 264 | 433 | 39.03 |
| **≤14** | Community | 718 | 1367 | 2085 | 34.44 |
| **>15** | Nosocomial | 165 | 258 | 423 | 39.01 |
| **≤15** | Community | 722 | 1373 | 2095 | 34.46 |

community-acquired covid -19.

Supplementary Figure 2: Kaplan Meier survival plot of nosocomial versus community infection of covid-19 patients.

Sensitivity analysis to investigate the performance of the commonly-used Kaplan Meier Survival Curve to compares survival in Community-acquired (light grey) and Nosocomial-acquired covid-19 infection (dark grey). Survival curves are compared using the log-rank test with 95% confidence interval indicated by shading. Here, censoring of patients into the absorbing state of “discharge” at a disproportionate rate within the Community-acquired covid-19 group leads to inverse interpretation to that seen in competing risks analysis.

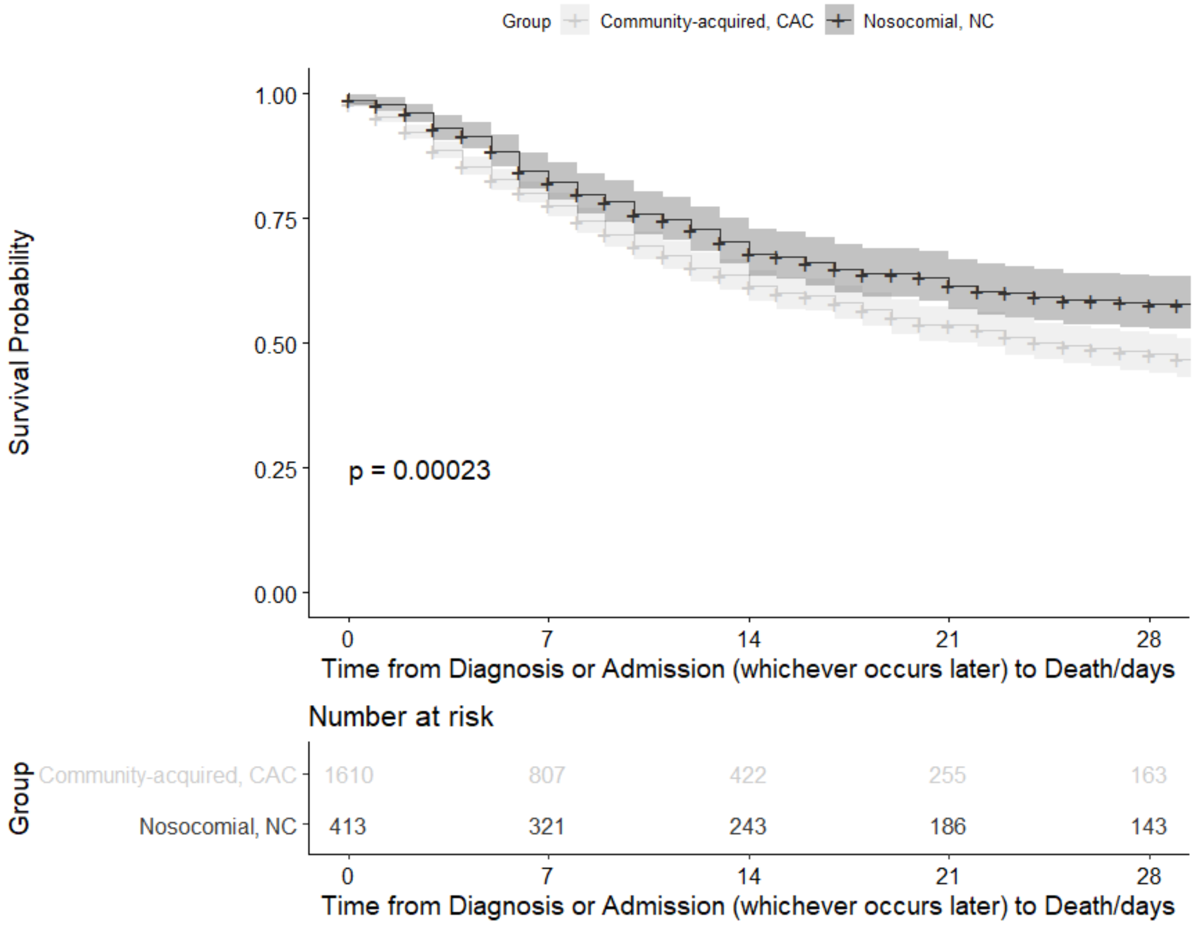

Supplementary Table 5: Comparison of community-acquired and nosocomial-acquired covid-19 patient characteristics, based on a diagnostic case testing threshold of 48 hours after admission

| **Variable, median (IQR)** | **Diagnosed within first 48 hours of admission "Community-Acquired"** | **Diagnosed after first 48 hours of admission "Nosocomial-Acquired"** | **Univariate Significance** |
|---|---|---|---|
| N | 1515 (60.2%) | 1003 (39.8%) | - |
| Female (n, %) | 663(43.8%) | 486(48.5%) | 0.0394 |
| Age, years | 69 (56-80) | 79 (71-87) | <0.0001 |
| Total co-morbidities count | 2.0 (1.0-4.0) | 3.0 (2.0-4.0) | <0.0001 |
| Clinical Frailty Score | 3 (2-6)<br>*Data available in 864 cases (57%)* | 6 (4-7)<br>*Data available in 445 cases (44%)* | <0.0001 |
| Welsh index of multiple deprivation | 745 (372-1283)<br>*Data available in 1442 cases (95%)* | 775 (403-1323)<br>*Data available in 952 cases (95%)* | 0.180 |

Supplementary Table 6: Monthly prevalence of nosocomial infection diagnosis

| **Month (2020)** | **Total recorded cases** | **COPE/ Public Health Wales case definition*** | **Clinician-recoded diagnosis** |
|---|---|---|---|
| | | 'Definite' nosocomial (%) | Hospital-acquired (%) |
| March | 681 | 97 (14.2%) | 118 (17.3%) |
| April | 1353 | 216 (16.0%) | 224 (16.6%) |
| May | 347 | 73 (21.0%) | 67 (19.3%) |
| June | 135 | 25 (18.5%) | 26 (19.3%) |

**Defined by diagnostic PCR testing performed >14 days following hospital admission.*
